# CLINICAL COURSE, RISK FACTORS FOR TRANSFER TO ICU AND MORTALITY IN PATIENTS WITH COVID-19 AFFECTED BY ACUTE RESPIRATORY FAILURE REFERRED TO A RESPIRATORY INTERMEDIATATE CARE UNIT

**DOI:** 10.1101/2020.08.19.20178350

**Authors:** Enrico Buonamico, Vitaliano Nicola Quaranta, Esterina Boniello, Michela Dimitri, Marco Majorano, Luciana Labate, Paola Pierucci, Elena Capozza, Giovanna Elisiana Carpagnano, Onofrio Resta

## Abstract

**Introduction:** There are no clear guidelines as yet for the selection of patients affected by COVID-19 who can be treated in intermediate RICU, neither shared criteria for their intubation and transfer in ICU.

In the present study we described the clinical course and risk factors for transfer to ICU and mortality of SARS-Cov-2 positive patients affected by acute respiratory failure, hospitalized in a Respiratory Intermediate Care Unit in the south of Italy.

**Methods:** In this retrospective, observational single centre study we evaluated 96 laboratory confirmed COVID-19 patients affected by acute respiratory failure (ARF). We compared demographic data, laboratory data and clinical outcomes between deceased and survived patients, aiming to identify risk factors for transfer to ICU and mortality, and possible gender-related differences.

**Results:** Of 96 patients, 51 (53.1%) survived and 45 (46.9 %) died. Among those who died, 23 (51.1%) deceased in RICU. Twenty-nine (30.2%) were transferred to ICU, of whom 22 (75.9%) died in ICU. Patients affected by COPD have a higher mortality compared to patients without this comorbidity (p = 0.002). Lower baseline P/F ratio (p = 0,014) and neurologic comorbidities (p = 0,008) emerged as risk factors for death.

Male were younger than female patients (66 vs 80 y.o.; p = 0.042). In female patients, lower peripheral blood lymphocyte count (p = 0.007) is a risk factor for death, characteristic gender-related in our sample.

Female sex was a protective parameter against transfer to ICU (p = 0,036) and P/F ratio wasn’t a significant predictor of transfer to ICU (p = 0,227).

Only higher baseline CRP (p = 0,034) has shown a predictive role for transfer to ICU in our sample. Patients deceased after a transfer to ICU had younger age (p = 0,000), lower median comorbidity number (p = 0,000), lower D-dimer (p = 0,029) and lower prevalence of female sex (p = 0,029).

**Discussion:** Mortality in our study was similar to that found in other studies involving patients in non-invasive ventilation. In our study older age and comorbidities play as predictors of death in COVID-19 patients. COPD, despite presenting low prevalence, is a risk factor for death, both in men and women. In female patients chronic ischemic heart disease and congestive heart failure are death predictors. High CRP and lymphopenia, linked to inflammatory status, are predictors of transfer to ICU. Patients transferred to ICU higher mortality than the others, and patients who die in ICU are mostly men, younger and have less comorbidities. Baseline P/F ratio is not a good predictor of transfer to ICU, while in our sample is a sensible predictor of death. More studies need to be performed on COVID-19 patients, in the urgency of COVID-19 pandemic persistence.

## INTRODUCTION

Coronavirus disease 19 (COVID-19) has been declared by the World Health Organization (WHO) as a public health emergency of international concern^1^. Isolated and described by Chinese scientists as a novel coronavirus (severe acute respiratory syndrome coronavirus 2, SARS-Cov-2), it was confirmed as the responsible for the COVID 19 pandemic in February 2020 by the WHO^2^. The clinical spectrum of COVID-19 infection appears to be wide, encompassing asymptomatic infection, mild upper respiratory tract illness and severe viral pneumonia with acute respiratory failure (ARF)^3^. Most of patients with moderate-severe respiratory acute syndrome and acute respiratory distress syndrome (ARDS) require hospitalization in intensive care unit (ICU). In China, among hospitalized patients with COVID-19, the percentage of patients who required ICU care has varied from 5% to 32%^4^.

Furthermore, during COVID-19 pandemic, invasive mechanical ventilation (IMV) was performed only in less than half of patients admitted in ICU, while all the other patients underwent NIV. Conversely in Italy, non-invasive respiratory support is delivered more often outside in ICU, in Respiratory Intermediate Care Units (RICU) by respiratory phusicians.

Unfortunately, there are no clear guidelines as yet for the selection of patients affected by COVID-19 who can be treated in intermediate RICU, neither shared criteria for their intubation and transfer in ICU.

## MATHERIALS AND METHODS

### Population

This is a retrospective, observational single centre study. We obtained the medical records and compiled data from 96 hospitalized adult inpatients that were in our intermediate RICU of Teaching Hospital “Policlinico” of Bari from March 11th 2020 to May 31th 2020. COVID-19 was diagnosed on the basis of the WHO interim guidance^5^. A confirmed case of COVID-19 was defined as a positive result on high-throughput sequencing or real time reverse trascriptase-polymerase chain reaction (RTPCR) assay of nasal and pharyngeal swab specimens. Only laboratory confirmed cases were hospitalized in our intermediate RICU and included in the analysis, and all of them were affected by acute respiratory failure (ARF).

### Data Collection

The demographic data, medical history, comorbidities and Charlson Comorbidity Index (in order to better evaluate comorbidity burden in every single patient), laboratory findings and respiratory parameters were collected within the first 12 hour following intermediate RICU admission; in addition we reported patients’ clinical outcomes that were evaluated as survived, deceased in RICU, transferred to ICU and deceased in ICU. The sample was collected according the STROBE Statement^6^. This study was approved by the Policlinico Hospital of University of Bari “Aldo Moro” institutional review board. Data were analyzed and interpreted by the authors.

### Statistical Analysis

We verified the distribution normality of continuous variables in our study through the Kolmogorov-Smirnov test. Continuous variables are expressed in mean +/− standard deviation (SD) if they presented normal distribution, otherwise are expressed as median (interquartile range). Nominal and dichotomous variables were expressed as percentage.

Asymmetrical continuous variables were compared with Mann-Whitney test, while those with a normal distribution were compared with Student T-test for independent variables.

Nominal variables were compared with Fisher Chi-square test and Mantel-Haenszel test.

We performed Lo-rank Mantel-Cox survival analysis with COPD patients.

We performed univariate and Cox multivariate survival analysis related to death event for all the parameters object of the present study.

Univariate and multivariate logistic regression analysis relative to the probability of transfer to ICU were performed.

For all the statistical analysis performed in the present study, we assumed a significance level of p<0.05.

Among the continuous variables in our study, BMI, P/F ratio and LDH have normal distribution; all the other continuous variables have an asymmetrical distribution.

## RESULTS

### General outcomes

General outcomes are summarized in Table 1: our sample consists of 96 consecutive patients, of whom 51 (53.1%) survived hospitalization, while 45 patients (46.9 %) died. Among those who died, 23 patients (51.1%) deceased in RICU. Twenty-nine patients (30.2%) were transferred to ICU, of whom 22 (75.9%) died in ICU.

**Table 1:** General Outcomes.

| Parameters | Number (%) |
| --- | --- |
| Sample patients | 96 |
| Survived | 51 (53.1 %) |
| Deceased (total) | 45 (46.9 %) |
| Deceased in RICU | 23 (51.1 %) |
| Transferred to ICU | 29 (30.2 %) |
| Deceased in ICU | 22 (75.9 %) |

### Comparison between deceased patients and survived patients

In Table 2 we described baseline values and comorbidities that are commonly considered in literature as possible risk factors for mortality in COVID-19.

**Table 2:** Comparison deceased/survived patients.

| Parameters | Deceased | Survived | Significance |
| --- | --- | --- | --- |
| Age<br>Median (IQR) | 79,0 (69,0 – 86,0) | 61,0 (57,0 – 72,0) | **0,000** |
| Sex % |  |  | 0,568 |
| F | 28,9 | 29,4 |  |
| M | 71,1 | 70,6 |  |
| CCI<br>Median (IQR) | 5 (4,0 – 7,5) | 3,0 (1,0 - 4,0) | **0,000** |
| Asthma | 2,3 | 4,4 | **0,517** |
| COPD | 34,9 | 4,4 | **0,000** |
| Interstitial lung disease | 2,4 | 2,4 | **0,747** |
| Hypertension | 88,6 | 61,7 | **0,002** |
| Renal failure | 68,2 | 28,9 | **0,000** |
| Neoplasm | 14,0 | 17,0 | **0,458** |
| Neurological disease | 34,9 | 4,3 | **0,000** |
| Diabetes mellitus type 2 | 27,9 | 19,1 | **0,232** |
| Congestive heart failure | 39,5 | 10,4 | **0,001** |
| Atrial Fibrillation | 18,6 | 4,2 | **0,030** |
| Previous acute myocardial infarction | 11,9 | 8,5 | **0,438** |
| Chronic ischemic heart disease | 42,5 | 17,0 | **0,004** |
| Dysthyroidism | 17,1 | 10,4 | **0,272** |
| Liver disease | 9,8 | 2,1 | **0,141** |
| Comorbidity number |  |  | **0,016** |
| 0 | 0,0 | 17,0 |  |
| 1 | 11,6 | 17,0 |  |
| 2 | 11,6 | 21,3 |  |
| 3 | 41,9 | 34,0 |  |
| 4 | 18,6 | 6,4 |  |
| 5 | 4,7 | 2,1 |  |
| 6 | 11,6 | 2,1 |  |
| P/F ratio<br>Mean ± DS | 189,00 ± 110,83 | 228,56 ± 70,60 | **0,000** |
| Peripheral blood lymphocyte<br>count<br>Median (IQR) | 705,97 (545,4 – 884,2) | 882,3 (647,8 – 1217,1) | **0,004** |
| LDH<br>Mean ± DS | 347,2 ± 127,5 | 318,45 ± 97,82 | 0,693 |
| CRP<br>Median (IQR) | 126,5 (80,4 – 163,7) | 82,8 (47,5 – 146,2) | **0,047** |
| D-dimer<br>Median (IQR) | 1360,0 (734,2 – 3126,5) | 933 (530,0 – 2280,0) | 0,090 |

We found that deceased patients had significantly older age, higher CCI, higher overall comorbidity number, lower baseline P/F ratio and blood lymphocyte count, higher baseline CRP.

In particular, among comorbidities, we verified that patients affected by COPD have a higher mortality compared to patients without this comorbidity (Fig. 1; Log-Rank (Mantel-Cox) Chi Square 9.274; p = 0.002).

**Fig. 1:**
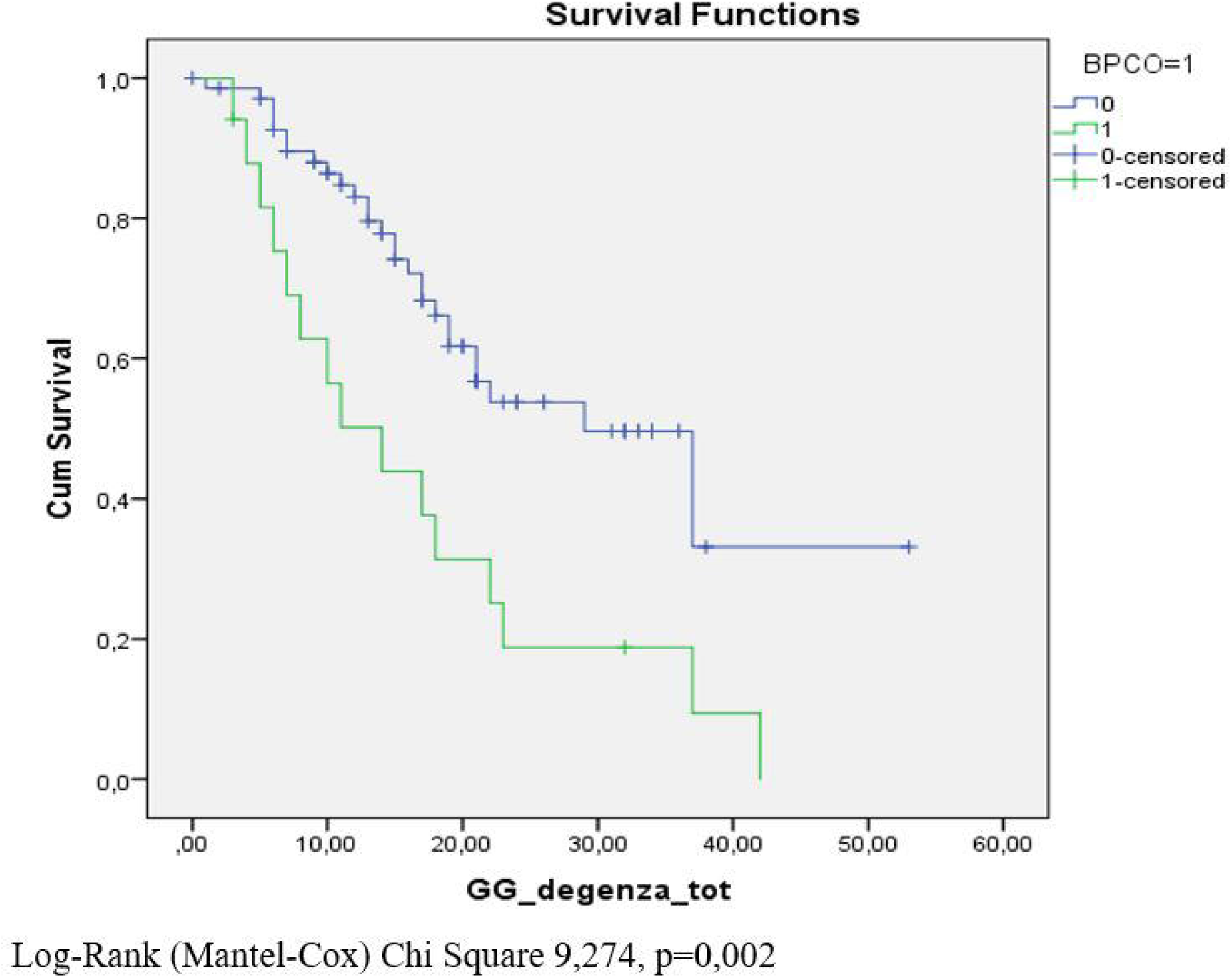
Survival curve relative to COPD comorbidity.

### Survival analysis

From the univariate analysis we found as risk factors for mortality older age (HR 1.049 (1.025 – 1.074); p = 0.000), a higher Charlson Index (HR 1,298 (1,161 – 1,452); p = 0,000), a low P/F ratio (HR 0,996 (0,992 – 0,999); p = 0,017), a low peripheral blood lymphocyte count (HR 0,999 (0,998 – 1,000); p = 0,035), COPD as comorbidity (HR 2,577 (1,355 – 4,900); p = 0,006) and congestive heart failure as comorbidity (HR 2,189 (1,169 – 4,096); p = 0,014). We found as mortality predictors also high levels of D-dimer, hypertension, renal failure and neurologic diseases as comorbidities.

Predictors found with univariate analysis were analysed using multivariate Cox analysis (Table 3), and lower baseline P/F ratio (HR 0,995 (0,991 – 0,999); p = 0,014) and neurologic comorbidities (HR 3,005 (1,342 – 6,957); p = 0,008) emerged as risk factors for death.

**Table 3.**
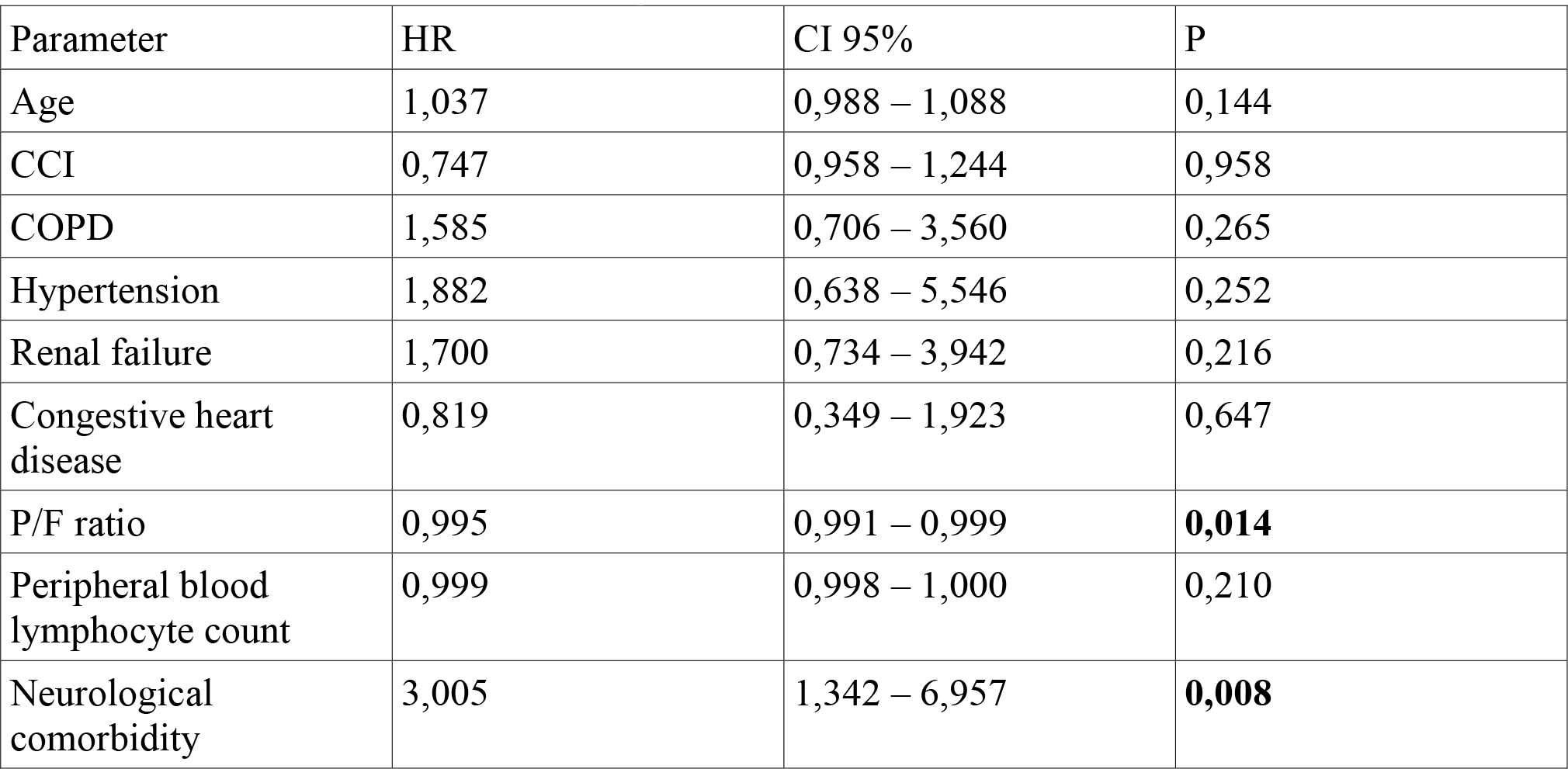
Multivariate Cox Analysis for probability of death.

| Parameter | HR | CI 95% | P |
| --- | --- | --- | --- |
| Age | 1,037 | 0,988 – 1,088 | 0,144 |
| CCI | 0,747 | 0,958 – 1,244 | 0,958 |
| COPD | 1,585 | 0,706 – 3,560 | 0,265 |
| Hypertension | 1,882 | 0,638 – 5,546 | 0,252 |
| Renal failure | 1,700 | 0,734 – 3,942 | 0,216 |
| Congestive heart<br>disease | 0,819 | 0,349 – 1,923 | 0,647 |
| P/F ratio | 0,995 | 0,991 – 0,999 | **0,014** |
| Peripheral blood<br>lymphocyte count | 0,999 | 0,998 – 1,000 | 0,210 |
| Neurological<br>comorbidity | 3,005 | 1,342 – 6,957 | **0,008** |

### Gender analysis

Male patients had significantly younger age compared to female patients (66 (58 – 77) vs 80 (65.25 – 88; p = 0.042). Male patients were divided in survived (36) and deceased (32) subgroups, and subgroups were compared for all parameters object of the study. Male deceased patients have significantly older age (71.93 +/− 13.5, median 72.5 vs 62.55 +/− 12.09 median 60.0; p = 0.001), higher CCI (median 5.0 (3.25 – 6.0) vs median 2.0 (1.0 – 4.0); p = 0000) and lower baseline P/F ratio (195.12 +/− 118.86; 184 (101.0 – 240.0) vs 232.75 +/−66.64; 232.0 (192.0 – 281.0); p = 0.017). Deceased patients presented a higher percentage of COPD (34.4% vs 6.5%; p = 0.006), hypertension (84.4% vs 56.3%; p = 0.014), renal failure (62.5% vs 22.6%; p = 0.001), neurological diseases (40.6% vs 3.1%; p = 0.000), congestive heart failure (31.3% vs 6.1%; p = 0.01), atrial fibrillation (18.8% vs 3.0%; p = 0.048). Comparison between subgroups relative to CRP, LDH, D-dimer and lymphocytes weren’t significant.

Similarly female patients were divided in survived (15) and deceased (13). Deceased female patients had significantly older age (84.30 +/−6.73 median 85 (80 – 89) vs 69.13 +/− 15.62 median 66.0 (57.0 – 83.0); p = 0.010), and, differently from male patients, lower peripheral blood lymphocyte count (665.17 +/− 635.44 median 635.06 (483.70 – 835.29) vs 1163.03 +/− 823.61 median 946.22 (836.0 – 1233.40); p = 0.007), characteristic that seems gender-related (Fig. 2). Moreover deceased patients had a higher prevalence of COPD (36.4% vs 0%; p = 0.026), renal failure (83.3% vs 42.9%; p = 0.042), congestive heart failure (63.6% vs 20%; p = 0.032). No difference regarding P/F ratio (p = 0.183), CCI, hypertension, neurologic comorbidity and atrial fibrillation between subgroups, differently from male patients. Differently from male patients, female deceased patients had higher prevalence of chronic ischemic heart disease compared to survived subgroup. Again, no difference was evidenced regarding LDH, CRP and D-dimer between subgroups also in female patients.

**Figure 2:**
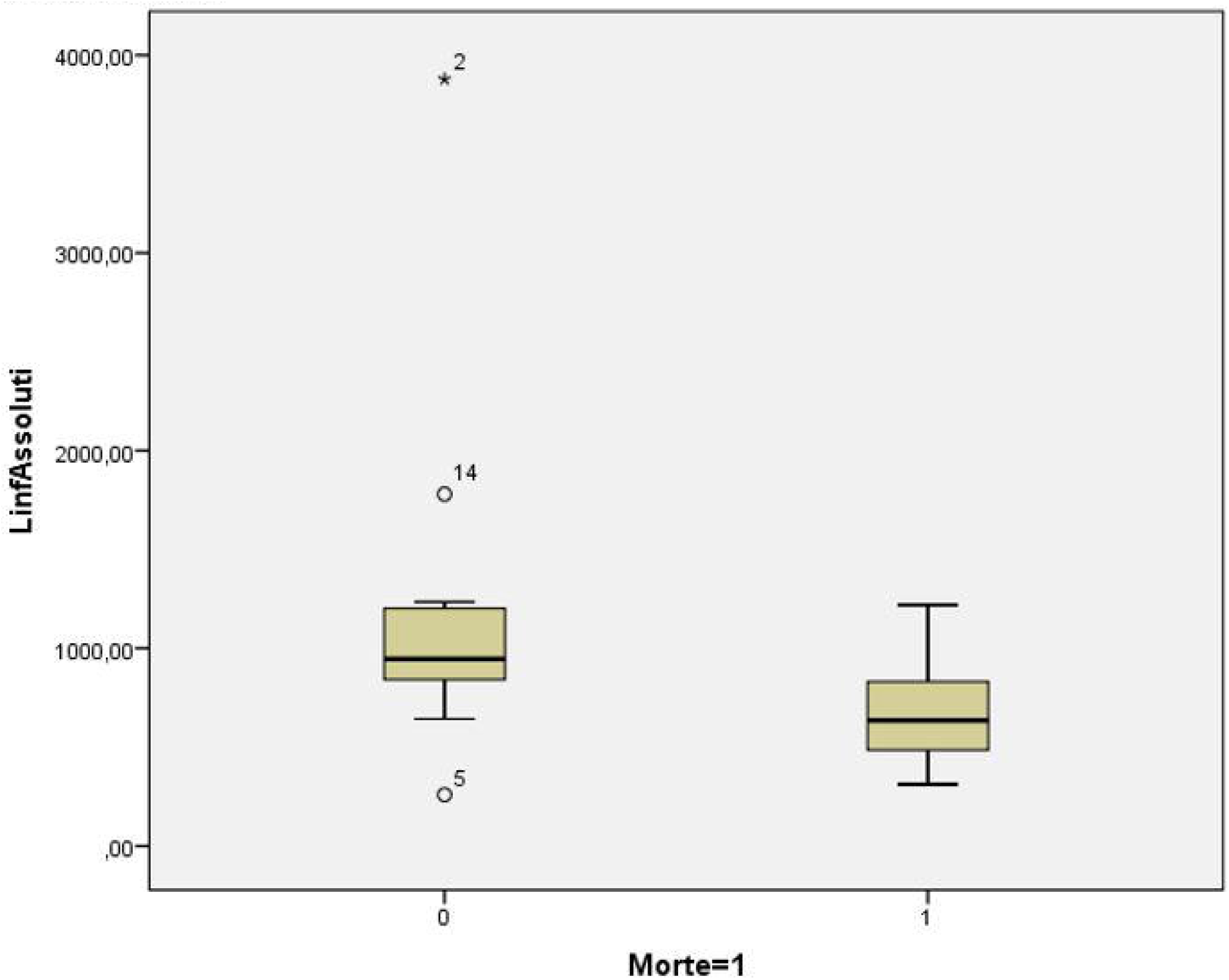
Box Plot. Comparison between deceased female patients (13) and survived female patients (15). Deceased patients had lower baseline blood lymphocyte count compared to survived (665,17±635,44 median 635,06 (483,70–835,29) vs 1163,03±823,61 median 946,22 (839,0–1233,40); p==0,007), that seems a gender-related characteristic, being not found in male patients.

### Probability of transfer to ICU

Twenty-nine patients (30.2%) were transferred to ICU. Using univariate linear regression analysis we verified that age was not a predictive parameter of transfer to ICU. Female sex was a protective parameter against transfer to ICU (O45/100*87R 0,287 [0,089 – 0,921]; p = 0,036), underlining once again the gender role in COVID-19 pathogenesis and evolution.

Patients transferred to ICU had 6.012 times the probability of death ([2,237 – 16,161]; p = 0,000).

P/F ratio was not significant in predicting the probability of transfer to ICU (OR 0,997; p = 0,227).

Factors raising significantly the probability of transfer to ICU were low lymphocyte count (OR 0,998 [0,997 – 1,000]; p = 0,038)) and higher baseline CRP values (OR 1,007 [1,001 – 1,013]; P = 0,018) (Table 5) →DIVENTA TAB 4.

**Table 4.** Univariate Logistic Regression for the probability of having ICU as final unit Parameter.

| Parameter | O.R. | CI 95% | p |
| --- | --- | --- | --- |
| Age | 0,987 | 0,957 – 1,018 | 0,410 |
| Female sex | 0,259 | 0,089 – 0,921 | **0,036** |
| BMI | 0,903 | 0,795 – 1,025 | 0,114 |
| CCI | 0,880 | 0,739 – 1,048 | 0,880 |
| P/F ratio | 0,997 | 0,992 -1,002 | 0,227 |
| Lymphocytes | 0,998 | 0,997 – 1,000 | **0,038** |
| CRP | 1,007 | 1,001 – 1,013 | **0,018** |

**Table 5.** Multivariate Logistic Regression for the probability of having ICU as final unit.

| Parameter | O.R. | CI 95% | p |
| --- | --- | --- | --- |
| Female sex | 0,316 | 0,091 -1,004 | 0,071 |
| Lymphocytes | 0,998 | 0,997 – 1,000 | 0,054 |
| CRP | 1,007 | 1,001 – 1,013 | **0,034** |

Parameters that significantly predict transfer to ICU obtained with univariate analysis (male sex, lymphopenia, high CRP) were tested with a multivariate model, underlining the importance of high CRP (OR 1,007[1,001 – 1,013]; p = 0,034) and lymphopenia that gets close to statistical significance (Table 6) → DIVENTA TAB 5.

**Table 6:** Comparison of subpopulation (transferred to ICU and not transferred to ICU) between deceased and survived.

| Subpopulation | Parametri | Deceased | Survived | P |
| --- | --- | --- | --- | --- |
|  | Numerosity | 7 | 22 |  |
| **Transferred to ICU (n=29)** | Age median (IQR) | 72,0 (64,7 – 76,7) | 60 (57,0 – 65,0) | 0,024 |
|  | CCI median (IQR) | 4,0 (2,7 – 5,2) | 2,0 (2,0 – 3,0) | 0,028 |
| **Not transferred to ICU (n=67)** | Numerosity | 23 | 44 |  |
|  | Age median (IQR) | 84,0 (77,5 – 89,0) | 61,0 (55,2 – 76,2) | 0,000 |
|  | CCI median (IQR) | 7,0 (5,0 – 9,0) | 3,0 (1,0 – 5,0) | 0,000 |
|  | Lymphocytes median (IQR) | 727,7 (482,4 – 915,2) | 852,3 (647,8 – 1217,2) | 0,028 |
|  | D-dimer median (IQR) | 1607,0 (888,0 – 5617,5) | 1100,5 (530,0 - 2792,5) | 0,039 |
|  | COPD % | 47,6 | 5 | 0,000 |
|  | Hypertension | 95,5 | 58,5 | 0,001 |
|  | Renal failure | 81,8 | 31,7 | 0,000 |
|  | Neurologic comorbidity | 38,1 | 2,4 | 0,000 |
|  | Congestive heart disease | 57,1 | 11,9 | 0,000 |
|  | Chronic ischemic heart disease | 61,9 | 14,3 | 0,000 |

### Subgroup analysis: comparison between deceased and survived patients transferred to ICU and deceased and survived patients not transferred in ICU

Twenty-two patients (75.9%) of 29 patients transferred to ICU died. Comparing in this subgroup deceased patients with survived, we found as determining factors age (72 [64,7–76,7] vs 60 [57,0 – 65,0]; p = 0,024) and Charlson index (4 [2,7 – 5,2] vs 2 [2,0 – 3,0]; p = 0,028) (Table 4) →DIVENTA TABELLA 6. No specific comorbidity was associated to death, suggesting that is the high comorbidity burden and not a specific comorbidity that is associated to death in ICU. All other parameters as P/F (p = 0.134) were not significant.

Among patients not transferred to ICU (n = 67), 23 (34.3%) died. Similarly to other subgroup older age (84,0 (77,5 – 89,0) vs 61,0 (55,2 – 76,2); p = 0,000) and higher CCI (7,0 (5,0 – 9,0) vs 3,0 (1,0 – 5,0); p = 0,000) were associated to death. Regarding comorbidity, deceased patients had significantly higher prevalence of COPD (47,6% vs 5%), hypertension, (95,5% vs 58,5%), renal failure (81,8% vs 31,7%), neurological comorbidities (38,1% vs 2,4%), congestive heart failure (51,1% vs 11,9%) and chronic ischemic heart failure (61,9% vs 14,3%).

Moreover low peripheral blood lymphocyte count (727,7 (482,4 – 915,2) vs 852,3 (647,8 – 1217,2); p = 0,028) and higher D-dimer values (1607,0 (888,0 – 5617,5) vs 1100,5 (530,0 – 2792,5); p = 0,039) were associated to death (Table 4) → DIVENTA TABELLA 6.

Again no difference emerged between subgroups regarding P/F ratio (201,0 (102,5 – 257,5) vs 222,0 (186,2 – 279,7); p>0,050)

### Sub-population analysis: comparison between deceased patients previously transferred to ICU and deceased patients not transferred to ICU

As shown in Tab. 7, patients deceased after a transfer to ICU had younger age (72,0 (64,5 – 75,0) vs 84,0 (77,5 – 89,0); p = 0,000), lower median comorbidity number (4 (2,5 – 5,5) vs 7, 0 (5,0 – 9,0); p = 0,000), lower D-dimer values (947,0 (637,5 – 2020) vs 1067,0 (888,0 –5617,5); p = 0,029) and lower prevalence of female sex (13.6% vs 43.5%; p = 0,029) (Table 7).

**Table 7.** Comparison between patients deceased in ICU and deceased not transferred to ICU.

|  | Transferred to ICU<br>(n=22) | Not transferred to ICU<br>(n=23) | p |
| --- | --- | --- | --- |
| Age<br>median (IQR) | 72,0 (64,5 – 75,0) | 84,0 (77,5 – 89,0) | 0,000 |
| CCI<br>median (IQR) | 4 (2,5 – 5,5) | 7 ,0 (5,0 – 9,0) | 0,000 |
| D-dimer<br>median (IQR) | 947,0 (637,5 - 2020) | 1067,0 (888,0 –5617,5) | 0,043 |
| Female sex % | 13,6 | 43,5 | 0,029 |

## DISCUSSION

Our population consists of 96 critical COVID-19 patients with mean age 69.65 +/− 14.33 years old, 28.9% of whom female, and 30.2% needed transfer to ICU, while 46.9% of the patients deceased.

In our study mortality, despite being considerable, must be compared with other experiences of COVID-19 units dedicated to non-invasive ventilatory support, and mortality rates in studies that consider non-invasive ventilation treated COVID-19 patients vary from 45% of this large multicentered German study^7^ to sensibly higher values (79%, 83.3%) of studies with smaller samples^8,9^.

Aim of our study was to identify mortality prognostic features, possible gender-related differences and to identify ICU transfer prognostic factors. Moreover we wanted to compare causes of death between patients transferred to ICU versus patients not transferred to ICU.

Deceased patients presented older age, a higher comorbidity number, lower baseline P/F. Moreover they had lower baseline blood lymphocyte count and higher baseline CRP.

Survival analysis confirmed the death predictive value of older age, lower baseline P/F ratio, lymphopenia, congestive heart failure. Also higher D-dimer, hypertension, renal failure and neurological comorbidity were found as factors predictive of death.

Moreover survival analysis evidenced that COVID-19 patients affected by COPD, pre-existing neurologic disease, hypertension or by chronic heart failure die significantly more than those without these comorbidity. About hypertension and chronic heart failure, this evidence follows several studies that link pre-existent cardiovascular diseases to higher mortality in COVID^10,11,12^, while results about association between COPD and COVID-19 are divergent: despite many studies evidence a low prevalence of COPD in COVID-19 patients^13,14^, the same COPD seems related to a higher (from 2 to 5 times) risk of developing a severe form of COVID-19^15,16,17^. In our experience, COPD plays an important role between all the comorbidities, raising mortality up to 250% (HR 2.577), with a solid statistical certainty obtained with a survival analysis. The interest into the association between COPD, the most frequent smoke-related disease and COVID-19 is due to the early and well demonstrated evidence of the up-regulation of ACE-2 in respiratory mucosa cells driven by the activation of α7 nAChR nicotinic receptor, which/h is, in turn, up-regulated by nicotinic exposure^18,19,20^. Despite this laboratory evidence, one of the possible explanations of this divergence is that a higher number of comorbidities is linked to a worse outcome in COVID-19 patients^11^, and COPD is by itself a comorbidity, furthermore linked to other comorbidities, especially metabolic and cardiovascular. However, waiting for further pathogenetic explanations of the role of COPD in COVID-19, an increased attention to COPD patients seems mandatory, obtaining a better pharmacologic control of the underlying comorbidity, especially through a better adherence of patients to inhaled therapy, and better strategies of contagion prevention.

The multivariate analysis emphasizes the role of neurologic comorbidity as a predictor of death in our patients. To our knowledge, there aren’t studies that investigate the link between pre-existing neurologic diseases and mortality in COVID-19 patients, but in our sample the higher mortality may be explained by the heavy comorbidity burden of these patients, mostly affected by Alzheimer disease and other neurodegenerative disorders, that is a well known mortality risk^11^.

Also baseline low P/F, as evidenced from multivariate analysis, is a sensible predictor of death in patients with COVID in our sample, and while this may not be surprising being low P/F one of the markers of severe community acquired pneumonia^21^ and above all, is the fundamental marker of ARDS according to the Berlin definition^22^, to the best of our knowledge this is the first study to evaluate baseline P/F as a risk factor for mortality in COVID-19. Part of current research on COVID-19 is focused on the predictive value of comorbidities, sex and age, but in our sample, independently from other factors, baseline P/F seems a reliable predictive factor, thus leading to underline the importance of respiratory failure in COVID-19, which has proven to be a multisystemic disease, but that targets the lung as the organ that suffers the most decisive damage.

A possible link between lower baseline P/F ratio and neurologic comorbidities may be due to the contemporary impairment of peripheral gas exchange and, likely, of the loss of central neurologic compensation of ventilatory drive, that cooperate in worsening the outcome of patients with these two features. Further studies that evaluate respiratory drive (for example with P0.1) are necessary to confirm this suggestion.

All our data concerning predictive factors for death confirm that the most our patients had markers of baseline criticality (older age, higher CCI) or disease criticality (lower P/F, higher CRP, lymphopenia), the higher is the association with mortality.

### Gender sub-analysis

To the best of our knowledge, there is a lack of studies that compare values and characteristics associated to mortality divided per gender. Both in male and female patients, dead patients had older age and higher prevalence of COPD and chronic kidney failure, but unexpectedly only in male patients higher CCI, hypertension, atrial fibrillation, pre-existing neurological diseases and lower baseline P/F were associated to death.

In female patients only chronic heart failure and chronic ischemic heart disease were associated to death. In particular, as shown in Fig. 1, a significantly lower blood lymphocytes count in dead patients seem a gender-related characteristic, as this association is shown only in female patients.

This interesting new evidence in our study is firm, but our sample is too small and we need further confirms to better understand the strength and the meaning of this difference.

The evidence of major baseline P/F ratio impairment as a factor associated to death in male patients only may be explained by the higher prevalence of chronic ischemic heart disease and chronic heart failure in women, leading to the suggestion in women that death was linked to heart failure rather than respiratory failure. Confirming this suggestion, female patients had also significantly older age, and suffered less transfer to ICU for acute respiratory failure unresponsive to non-invasive ventilation.

### Probability of transfer to ICU

To our knowledge there aren’t study that explore risk factors for transfer to ICU in COVID-19 patients. In our sample only high CRP and low blood lymphocyte count showed a predictive role for transfer to ICU, while female sex showed a protective role against transfer to ICU. Baseline P/F ratio role as a risk factor for negative outcome is debunked when considered for transfer to ICU. In our experience, P/F ratio suffered a high variability throughout patient’s hospitalization, often showing rapid falls from baseline, needing sudden patient’s intubation and transfer to ICU.

Differently laboratory parameters of disease severity, such as CRP and lymphocytes count, worked as risk factors from baseline, suffering less variability until any improvement in clinical conditions.

### Role of mortality associated to transfer to ICU

Patients transferred to ICU suffer higher mortality than the others, in fact they are 6.012 times more likely to die. We further observed that among deceased patients, those who died in ICU had lower median age (72 vs 84), lower median comorbidity number (4 vs 7) and D-Dimer lower mean values (947 vs 1067) but unexpectedly not a significantly different P/F ratio, showing baseline similar respiratory condition. The explanation of these findings are linked to the emergency condition of COVID-19 pandemic: ICU beds were reserved to, in case of similar critical conditions, younger and less comorbid patients, for this reason patients who remained in RICU were older and affected by a higher comorbidity burden. The higher mortality rate of patients transferred to ICU may be explained, conversely, by common ICU mortality risks, such as intubation and antibiotic resistant bacterial super-infections. Another possible explanation is a more powerful cytokine storm in younger patients, associated to higher mortality risk, but further studies need to be conducted in order to deepen our knowledge of mortality causes in younger COVID-19 patients. In our sample patients deceased in ICU show a lower prevalence of female sex compared to patients deceased outside ICU, and this evidence may be explained by the older age of women in our population, that once again is a factor related against transfer to ICU.

## CONCLUSION

Our study confirms that pre-existing factors that are older age and comorbidities play an important role in determining disease mortality in COVID-19 patients. COPD, though being a low-prevalence comorbidity in COVID-19 patients, gives patients a more severe disease course conditioning the outcome, both in men and women. In female patients chronic ischemic heart disease and congestive heart failure are death predictors. Predictors of transfer to ICU (high baseline CRP and lymphopenia) are related more to inflammatory status that influences disease severity rather than age and comorbidities. Patients transferred to ICU suffer a far higher mortality than those who are not transferred, and patients who die in ICU are mostly men, but are unexpectedly younger and have few comorbidities, this due.

P/F ratio is the most important marker of ARDS, but in baseline is not a good predictor of transfer to ICU, probably because of the trend of COVID-19 patients, that suffer sudden worsening in respiratory failure. On the contrary baseline P/F in our sample is a very sensible predictor of death. More studies need to be performed on COVID-19 patients, including larger samples, in order to identify as precisely as possible risk factors for severe disease and for death, in the urgency of a pandemic that, after more than 8 months from the very beginning, still puts to a tough test health systems all over the world.

## Data Availability

All data referred to the manuscript are available.

## Bibliography

1. World Health Organization. WHO Director-General’s opening remarks at the media briefing on COVID-19 – 11 March 2020. 2020;(March).

2. Wang C, Horby PW, Hayden FG, Gao GF. A novel coronavirus outbreak of global health concern. Lancet. 2020;395:15–18. doi:10.1016/S0140-6736(20)30185-9

3. Zhou F, Yu T, Du R, et al. Clinical course and risk factors for mortality of adult inpatients with COVID-19 in Wuhan, China : a retrospective cohort study. Lancet. 2020;395(10229):1054–1062. doi:10.1016/S0140-6736(20)30566-3

4. Guan W, Ni Z, Hu Y, et al. Clinical Characteristics of Coronavirus Disease 2019 in China. 2020;382:1708–1720. doi:10.1056/NEJMoa2002032

5. World Health Organization. Public health surveillance for COVID-19. 2020.

6. von Elm E, Altman DG, Egger M, Pocock SJ, Gøtzsche PC, Vandenbroucke JP. The Strengthening the Reporting of Observational Studies in Epidemiology (STROBE) statement : guidelines for reporting observational studies. Lancet. 2007;370:1453–1457. doi:https://doi.org/10.1016/S0140-6736(07)61602-X

7. Karagiannidis C, Mostert C, Hentschker C, et al. Case characteristics, resource use, and outcomes of 10 021 patients with COVID-19 admitted to 920 German hospitals: an observational study. Lancet Respir Med. 2020;(2600):1–10. doi:10.1016/S2213-2600(20)30316-7

8. Yang X, Yu Y, Xu J, et al. Clinical course and outcomes of critically ill patients with SARSCoV-2 pneumonia in Wuhan, China: a single-centered, retrospective, observational study. Lancet Respir Med. 2020;8(5):475–481. doi:10.1016/S2213-2600(20)30079-5

9. Sivaloganathan AA, Nasim-mohi M, Brown MM, et al. Noninvasive ventilation for COVID-19 associated acute hypoxaemic respiratory failure: experience from a single centre. Br J Anaesth. 2020. doi:10.1016/j.bja.2020.07.008

10. Gao J, Wu F. Association between fractional exhaled nitric oxide, sputum induction and peripheral blood eosinophil in uncontrolled asthma. Allergy, Asthma Clin Immunol. 2018:1–9. doi:https://doi.org/10.1186/s13223-018-0248-7

11. Guan W, Liang W, Zhao Y, et al. Comorbidity and its impact on 1590 patients with COVID-19 in China : a nationwide analysis. Eur Respir J. 2020;55(March 2020):1–14. doi:10.1183/13993003.00547-2020

12. Wang D, Hu B, Hu C, et al. Clinical Characteristics of 138 Hospitalized Patients With 2019 Novel Coronavirus–Infected Pneumonia in Wuhan, China. JAMA. 2020;323(11):1061–1069. doi:10.1001/jama.2020.1585

13. Rossato M, Russo L, Mazzocut S, Vincenzo A Di, Fioretto P, Vettor R. Current Smoking is Not Associated with COVID-19. Eur Respir J. 2020;55:4–7.

14. Farsalinos K, Angelopoulou A, Alexandris N, Poulas K. COVID-19 and the nicotinic cholinergic system. Eur Respir J. 2020;56:2–5. doi:10.1183/13993003.01589-2020

15. Wang B, Li R, Lu Z, Huang Y. Does comorbidity increase the risk of patients with COVID-19 : evidence from meta-analysis. Aging (Albany NY). 2020;12(7):6049–6057. doi:10.18632/aging.103000

16. Zhao Q, Meng M, Lian N, et al. The impact of COPD and smoking history on the severity of COVID‐19 : A systemic review and meta‐analysis. J Med Virol. 2020;April:1–7. doi:10.1002/jmv.25889

17. Lippi G, Henry BM. Chronic obstructive pulmonary disease is associated with severe coronavirus disease 2019 (COVID-19). Respir Med. 2020;167(January):1–2. doi:10.1016/j.rmed.2020.105941

18. Zhou P, Yang X, Wang X, et al. A pneumonia outbreak associated with a new coronavirus of probable bat origin. Nature. 2020;579(January):270–273. doi:10.1038/s41586-020-2012-7

19. Leung JM, Yang CX, Sin DD. Reply to: “Current smoking is not associated with COVID-19”. Eur Respir J. 2020;55:2–3. doi:10.1183/13993003.01340-2020

20. Russo P, Bonassi m S, Giacconi R, Malavolta M, Tomino C, Maggi F. COVID-19 and smoking: is nicotine the hidden link? Eur Respir J. 2020;55:1–3. doi:10.1183/13993003.01116-2020

21. Metlay JP, Waterer GW, Long AC, et al. AMERICAN THORACIC SOCIETY Diagnosis and Treatment of Adults with Community-acquired Pneumonia An Official Clinical Practice Guideline of the American Thoracic Society and Infectious Diseases Society of America. Am J Crit Care Med. 2019;200(7):45–67. doi:10.1164/rccm.201908-1581ST

22. The ARDS Task Force. Acute Respiratory Distress Syndrome. The Berlin Definition. JAMA. 2012;307:2526–2533. doi:10.1001/jama.2012.5669

